# Proteomic aging clock (PAC) predicts age-related outcomes in middle-aged and older adults

**DOI:** 10.1101/2023.12.19.23300228

**Authors:** Chia-Ling Kuo, Zhiduo Chen, Peiran Liu, Luke C. Pilling, Janice L. Atkins, Richard H. Fortinsky, George A. Kuchel, Breno S. Diniz

## Abstract

Beyond mere prognostication, optimal biomarkers of aging provide insights into qualitative and quantitative features of biological aging and might, therefore, offer useful information for the testing and, ultimately, clinical use of gerotherapeutics. We aimed to develop a proteomic aging clock (PAC) for all-cause mortality risk as a proxy of biological age. Data were from the UK Biobank Pharma Proteomics Project, including 53,021 participants aged between 39 and 70 years and 2,923 plasma proteins assessed using the Olink Explore 3072 assay®. The Spearman correlation between PAC proteomic age and chronological age was 0.77. A total of 10.9% of the participants died during a mean follow-up of 13.3 years, with the mean age at death 70.1 years. We developed a proteomic aging clock (PAC) for all-cause mortality risk as a surrogate of BA using a combination of least absolute shrinkage and selection operator (LASSO) penalized Cox regression and Gompertz proportional hazards models. PAC showed robust age-adjusted associations and predictions for all-cause mortality and the onset of various diseases in general and disease-free participants. The proteins associated with PAC were enriched in several processes related to the hallmarks of biological aging. Our results expand previous findings by showing that age acceleration, based on PAC, strongly predicts all-cause mortality and several incident disease outcomes. Particularly, it facilitates the evaluation of risk for multiple conditions in a disease-free population, thereby, contributing to the prevention of initial diseases, which vary among individuals and may subsequently lead to additional comorbidities.

## 1 Introduction

The geroscience hypothesis (Kennedy et al. 2014) posits that targeting aging may prevent or delay the onset of multiple diseases, where chronological age is a major risk factor. Human trials to test interventions for a geroscience indication are challenging due to the lengthy duration needed to observe the effects of interventions on aging outcomes. Alternatively, biomarkers of aging, reflective of aging processes and their consequences, can serve as surrogate endpoints for assessing the risk and progression of several major diseases (Justice et al. 2018; Moqri et al. 2023). Recent biomarkers of aging focus on prediction of biological age (BA) (Rutledge et al. 2022; Moqri et al. 2023), which reflects the level of age-dependent biological changes, such as molecular and cellular damage accumulation (Moqri et al. 2023). Biological age (BA) acceleration, indicated by the difference between BA and chronological age, has been linked with various adverse health outcomes (Rutledge et al. 2022; Moqri et al. 2023). Due to their prognostic value in predicting age-related conditions and sensitivity to interventions, BA measures have become increasingly popular as intermediate phenotypes in randomized controlled trials (Waziry et al. 2023; Lohman et al. 2023).

The most common quantification of BA relies on DNA methylation-based measurements known as epigenetic clocks. First-generation epigenetic clocks (Hannum et al. 2013; Horvath 2013; Levine 2013, p.201) were initially developed using chronological age as a surrogate of BA. These measures are now being surpassed by second-generation epigenetic clocks, which use an age-related outcome or the pace of aging to improve predictions of morbidity and mortality (Levine et al. 2018; Lu et al. 2019; Belsky et al. 2022). While epigenetic clocks are widely recognized, proteins and their signatures provide direct links to aging-related pathology, making them more relevant for disease prognosis in the clinical context (Moaddel et al. 2021).

Previous studies have identified circulating proteins associated with chronological age (Moaddel et al. 2021; Sathyan et al. 2020; Tanaka et al. 2018), mortality (Orwoll et al. 2018; Eiriksdottir et al. 2021), and chronic diseases (Gadd et al. 2023; Carrasco-Zanini et al. 2023). Several proteomic clocks have been developed to predict chronological age (Tanaka et al. 2018; Sayed et al. 2021; Sathyan et al. 2020) or mortality (Unterhuber et al. 2021). In addition, a proteomic composite, known as the senescence-associated secretory phenotype (SASP) index (Diniz et al. 2017), was developed incorporating 22 pre-selected SASP proteins. This index indicates a phenotypic manifestation of cellular senescence, a hallmark of biological aging (López-Otín et al. 2013). Elevated SASP index levels have been associated with aging outcomes in older adults with major depression, such as cognitive impairment, increased medical burden– particularly in cardiovascular disease–and compromised brain health, including neuroinflammation and cortical atrophy (Seitz-Holland et al. 2023). Consistently, other composite SASP biomarkers have shown high predictive value for all-cause mortality in healthy older adults (St Sauver et al. 2023).

Despite their relevance, these proteomic clocks exhibit significant limitations. They were mostly trained to predict chronological age (i.e., first-generation clocks) or trained in a small sample with a small set of proteins, which may be pre-selected to reflect a specific biological aging process (e.g., cellular senescence). In this study, we aimed to develop a proteomic clock, referred to as the proteomic aging clock (PAC), to predict all-cause mortality. Data included 2,923 plasma proteins, assessed using the Olink Explore 3072 assay^®^, from a UK Biobank (UKB) baseline cohort of 53,021 participants aged between 39 and 70 years. We validated the PAC for its associations and predictions of all-cause mortality and age-related conditions, during a follow-up exceeding a decade. Biological insights into PAC proteomic age acceleration were investigated through gene set analysis and gene property analysis for tissue specificity.

## 2 Methods

### 2.1 UK Biobank Pharma Proteomics Project

The UK Biobank (UKB) recruited more than 500,000 participants, aged between 40 and 70 years, between 2006 and 2010 (Sudlow et al. 2015; Allen et al. 2024). At recruitment (baseline), participants completed online questionnaires, cognitive function tests, verbal interviews, and physical measurements. Additionally, blood samples were collected for future biological assays. Since then, disease diagnoses and death status have been updated through linkages to electronic health records.

Participants who supplied blood samples at baseline were selected for inclusion in the UK Biobank Pharma Proteomics Project (UKB-PPP) (Sun et al. 2023). Of the included samples (n=53,021), the majority (n=46,792, 88.3%) were a random sample from the UKB baseline cohort. Others (n=6,229, 11.7%) included participants who attended the first imaging visit and COVID-19 repeat imaging study and those selected by the consortium of 13 biopharmaceutical companies for their research interests.

### 2.2 Data

The normalized protein expression (NPX) data encompassed 2,923 proteins (**Table S1**). Three proteins with high rates of missing data were removed from the analysis: GLIPR1 (99.7%), NPM1 (74.0%), and PCOLCE (63.6%). For the remaining proteins, we applied a *k*-nearest neighbors approach (Torgo 2011) to impute missing proteomic data (*k*=10).

All-cause mortality risk was used as an indicator for BA. Death data were provided by the UK National Health Service (NHS) England, NHS Central Register, and National Records of Scotland. Participants with no recorded date of death were assumed to remain alive until the censoring date of 11/30/2022. Participants free of the disease at baseline (2006-2010) were followed up until the first disease diagnosis, death (censoring date 11/30/2022), or the last follow-up date (censoring dates: 11/30/2022 [England], 7/31/2021 [Scotland], 2/28/2018 [Wales]) depending on which occurred first. First diagnosis dates were identified using the UKB hospital inpatient data and first occurrence data, which linked data from different sources based on 3-character ICD-10 codes: longitudinal primary care (45% of the UK Biobank cohort), hospital inpatient, death registry data, and self-reported medical conditions at baseline (**Table S2**). Data on the covariates were collected by UKB through online surveys, physical measurements, and linkages to electronic health records (**Table S2**).

### 2.3 PAC development

The NPX data and chronological age at baseline in the training set were used to train a LASSO penalized Cox regression model for the risk of all-cause mortality. The selected proteins and chronological age were used to fit Gompertz proportional hazards models and formulate PAC to estimate the proteomic age based on the input data (**Supplementary Methods**).

### 2.4 PAC validation

#### 2.4.1 Correlations of PAC proteomic age with aging-related traits at baseline

We evaluated the correlations of PAC proteomic age with chronological age, BioAge, PhenoAge, short leukocyte telomere length (LTL), physiological or cognitive measures (**Table S3**), a 49-item frailty (Williams et al. 2019), and disease-associated biomarkers (**Table S3**) – all measured at baseline – using the test set data. Additionally, we investigated the correlations between the residuals of PAC proteomic age and those of BioAge, PhenoAge, and leukocyte telomere length, with adjustments made for chronological age.

#### 2.4.2 Associations of PAC proteomic age deviation with all-cause mortality and incident diseases

Next, we tested if PAC proteomic age deviation was linked with mortality and incident diseases (hypertension, myocardial infarction, heart failure, stroke, type 2 diabetes, COPD, pneumonia, chronic kidney disease, dementia, delirium, Parkinson’s disease, any cancer excluding non-melanoma skin cancer, and common cancers including breast cancer [females only], prostate cancer [males only], lung cancer, and colorectal cancer).

Using the test set data, we applied Cox regression models for all-cause mortality and Fine-Gray subdistribution hazard models for incident diseases to account for the effect of death. The models above were adjusted for each of the three sets of covariates at baseline: 1) **age-adjusted models**: age only, 2) **partially adjusted models**: sociodemographic factors (age, self-reported sex, ethnicity [White, Black, Asian, Other], and education [from none to college or university degree], Townsend deprivation index [higher values indicating higher levels of material deprivation]) and lifestyle factors (smoking status [current, former, never], body mass index [BMI]), and 3) **fully adjusted models**: covariates in the partially adjusted model and pre-existing diseases (hypertension, myocardial infarction, heart failure, stroke, type 2 diabetes, chronic obstructive pulmonary disease [COPD], pneumonia, chronic kidney disease, dementia, delirium, Parkinson’s disease, any cancer excluding non-melanoma skin cancer).

We also carried out a subgroup analysis stratifying the sample by sex. The p-values from the age-adjusted, partially adjusted, and fully adjusted models for all-cause mortality and incident diseases were adjusted for multiple testing using the Benjamini-Hochberg false discovery rate (FDR) method.

#### 2.4.3 PAC versus other BA measures in associations of biological age deviation with all-cause mortality and incident diseases

Using the test set data, the associations of PAC proteomic age with all-cause mortality and incident diseases were compared with those of other BA measures, namely BioAge, PhenoAge, and LTL, adjusting for the full set of covariates. BioAge (Levine 2013) was trained for chronological age, while PhenoAge (Levine et al. 2018) was trained for all-cause mortality, both using routine clinical biomarkers from blood samples in the National Health and Nutrition Survey (NHANES) III (detailed in **Supplementary Method**s). Further validation of both measures was performed in additional cohorts, including UKB (Liu et al. 2018; Kuo et al. 2021), confirming their robustness. LTL was assessed using a multiplex qPCR technique as T/S ratio, which compares the telomere amplification product (T) to that of a single-copy gene (S), adjusting for technical parameters (Codd et al. 2022). The rank-based inverse normal transformation was applied to each BA measure to convert the data to z-scores to standardize the scales of different BA measures. Short LTL by reversing the signs of LTL was compared with other BA measures so the association direction tended to be consistent across measures.

For sensitivity analysis, the associations above were investigated in participants without any pre-existing diseases at baseline. The primary fully adjusted models were simplified to the partially adjusted models as none of the included participants had developed any of the diseases.

#### 2.4.4 PAC versus other BA measures in predictions for all-cause mortality and incident diseases

Harrell’s C-statistic, a concordance probability within the range from 0.5 to 1, compares individuals in a pair that the individual who has a shorter time to a disease also has a higher risk for the disease based on the model during the follow-up time. Harrell’s C-statistic serves as a standard output to quantify discriminative power for Cox regression models, yet it demands an extended computation time for Fine-Gray subdistribution hazard models. Although we used Fine-Gray subdistribution hazard models to link BA deviation with incident diseases to account for the competing event of death, corresponding Cox regression models, which censored individuals who died before disease diagnosis yielded similar associations (results not shown). Without losing the generalization of our findings, we opted for Cox regression models to assess the predictions of PAC proteomic age against other BA estimates for all-cause mortality and incident diseases using the test set data.

#### 2.4.5 Functional analysis

To unravel the biological processes underlying BA deviation, proteins after the inverse normal transformation were associated with PAC proteomic age, BioAge, PhenoAge or short LTL in the fully adjusted linear regression models using the test set data. Significant proteins (Bonferron-corrected p<0.05) were carried forward to perform a gene set analysis and a gene property analysis for tissue specificity using the Functional Mapping and Annotation of Genome-Wide Association Studies (FUMA version 1.6.0) (Watanabe et al. 2017). Similar analyses were conducted for BioAge, PhenoAge, and short LTL.

In the gene set analysis, genes associated with BA deviation were compared with the background genes (20,260 protein-coding genes) for the presence in a hallmark gene set using a hypergeometric test. Enriched hallmark gene sets with at least five genes overlapped with the input genes were identified at the Bonferroni-corrected level of 5% (50 hallmark gene sets in total).

In the gene property analysis for tissue specificity, the input genes were compared with the background genes (protein-coding genes with mean normalized log_2_ expression value > 1 in at least one of 30 general tissues) using a hypergeometric test for the presence in a tissue-specific differentially expressed gene set (genes with p-value ≤ 0.05 after Bonferroni correction and absolute log fold change ≥ 0.58 in GTEx v8). Bonferroni-corrected p-values smaller than 5% were considered statistically significant.

## 3. Results

### 3.1 Training and test datasets

Participants with complete NPX and chronological age data from baseline (recruitment), and mortality data throughout the follow-up period were included in the PAC development. The included samples (n=53,021) were split into a training set (70%, n=37,115) and a test set (30%, n=15,906). In the training set, 4,034 participants (10.9%) died at the mean age of 70.1 years (SD=8.1) over a mean follow-up of 13.3 years (SD=2.2). Within the test set, 1,731 participants (10.9%) died, with the mean age at death 70.1 years (SD=8.1) during a mean follow-up of 13.3 years (SD=2.2). A baseline summary for participants in the training and test sets versus others in the UKB baseline cohort is presented in **Table S3**. Data were extracted using the field IDs in **Table S2**. The training and test samples showed comparable baseline characteristics to the rest of the UKB baseline cohort (**Table S3**). The disease prevalence was slightly higher within the UKB-PPP than the rest of the baseline cohort, which is expected, due to the enrichment of diseases in the UKB-PPP samples (Sun et al. 2023).

### 3.2 Development of the proteomic aging clock (PAC)

Using the training set data, a Least Absolute Shrinkage and Selection Operator (LASSO) penalized Cox regression model was applied to 2,920 proteins and chronological age at baseline to predict the time-to-event outcome of death. Chronological age and 128 proteins remained in the model (lambda 0.004543) (**Table S4**) and were carried forward to fit a Gompertz model.

Additionally, another Gompertz model was fitted to predict death solely using chronological age. We calculated the PAC proteomic age based on the shape and rate parameters, and the regression coefficients associated with individual proteins from the models above (**Table S5**). The mean PAC proteomic age was 53.4 years, 3.4 years younger than the mean chronological age in the training set. In the test set sample, the mean PAC proteomic age and chronological age were 53.4 and 56.9 years, respectively.

### 3.3 Correlations between chronological age, PAC proteomic age, PhenoAge, BioAge, LTL, plus a selection of aging phenotypes at baseline

A total of 10,451 participants had complete data for chronological age, PAC proteomic age, BioAge, PhenoAge, and LTL in the test set. The Spearman correlation (T) between PAC proteomic age and chronological age was 0.77, lower than the correlations of BioAge (T=0.98) and PhenoAge (T=0.87) with chronological age (**Figure S1**). Short LTL demonstrated weak correlations with chronological age and other BA measures (T≈0.2) (**Figure S1**). Single physiological or cognitive measures, frailty, and disease-associated biomarkers showed a weak to moderate association with chronological age, PAC proteomic age, BioAge, and PhenoAge (**Figures S1 and S2**). Additionally, the correlations between the age-adjusted residuals of PAC proteomic age and those of BioAge (*r*=0.07), PhenoAge (*r*=0.37), and LTL (*r*= -0.12), varied from low to moderate. This suggests that these biological age measures may represent distinct facets of biological aging.

### 3.4 Associations of PAC proteomic age acceleration with all-cause mortality and incident diseases

The PAC proteomic age acceleration showed significant associations with all-cause mortality and various incident diseases in the test set sample adjusting for chronological age only (age-adjusted model). These associations were attenuated in the partially adjusted (sociodemographic factors including age and lifestyle factors) and fully adjusted models (covariates in the partially adjusted model and pre-existing diseases), though remaining statistically significant (**Figure 1; Table S6**). For instance, the HR for all-cause mortality was 1.097 per year increase in PAC proteomic age (95% CI 1.091 to 1.103, p_adj_=3.83×10^-232^) in the fully adjusted model, versus 1.104 in the age-adjusted model, and 1.102 in the partially adjusted model. For sensitivity analysis, we included an indicator for participants selected by the UKB-PPP consortium in the fully adjusted models. The indicator was assigned a value of 1 for individuals chosen by the consortium due to specific diseases of interest and 0 for those not chosen by the consortium, selected randomly from the UK Biobank baseline cohort. The results were similar to the fully adjusted model results, likely attributable to the overlap in effects between the selection and baseline disease states. Similar results were also found in males and females separately (**Figures S3 and S4**).

**Figure 1.**
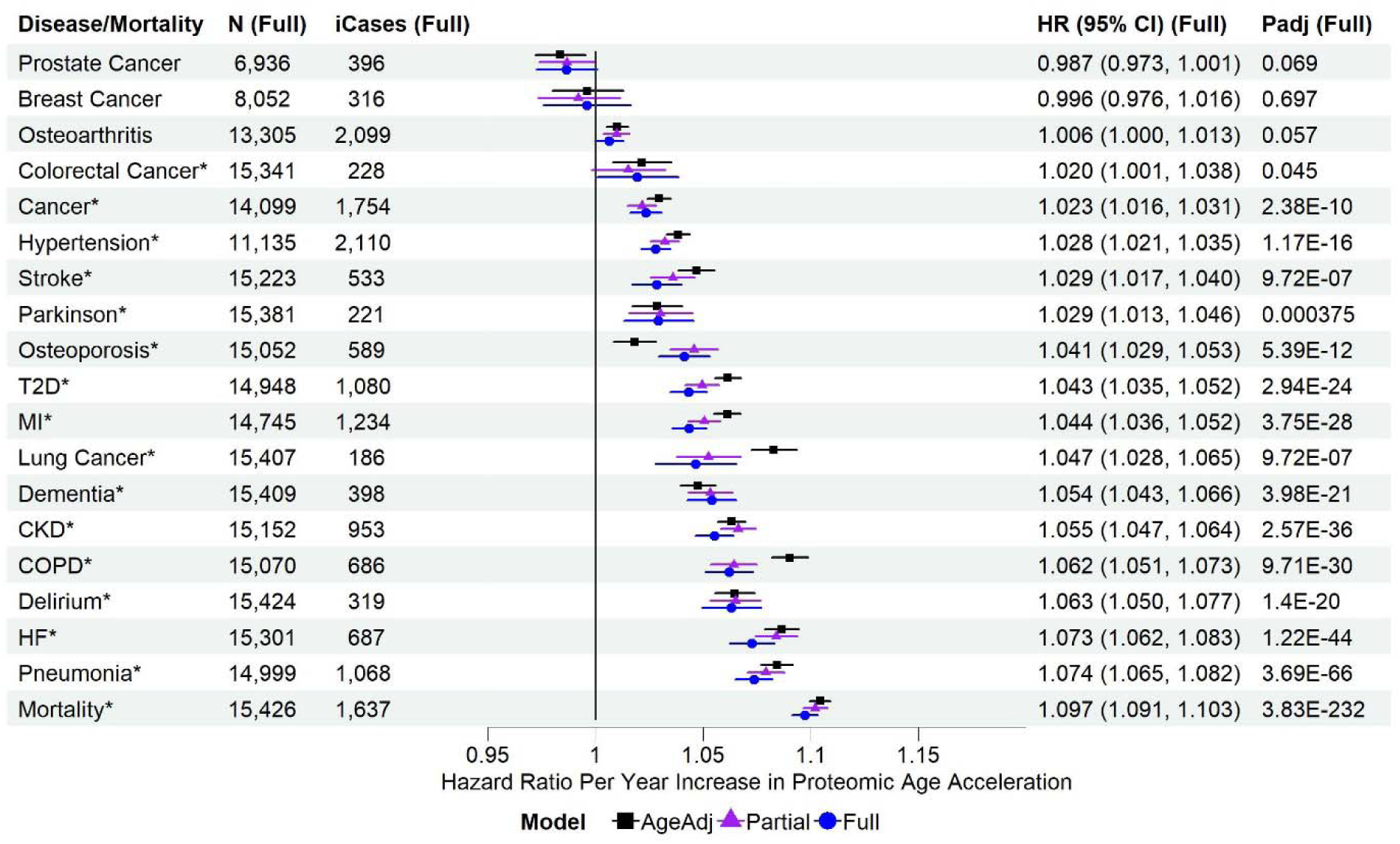
Associations of PAC proteomic age acceleration with all-cause mortality and incident diseases in the test set sample. MI: myocardial infarction; T2D: type 2 diabetes; COPD: chronic obstructive pulmonary disease; CKD: chronic kidney disease; HF: heart failure. N (Full): sample size with complete data for the fully adjusted model, after excluding participants diagnosed with the disease at or prior to baseline. iCases (Full): number of incident cases of N samples. Cox regression model for all-cause mortality and Fine-Gray sub-distribution hazard models to account for the effect of death on the risk for incident diseases, adjusting for different sets of covariates at baseline (age-adjusted, partially adjusted, and fully adjusted models). AgeAdj: chronological age; Partial: chronological age, sex, ethnicity, education, Townsend deprivation index, smoking status, and body mass index; Full: covariates in the partially adjusted model, and pre-existing diseases (hypertension, myocardial infarction, heart failure, stroke, type 2 diabetes, COPD, pneumonia, chronic kidney disease, any cancer excluding non-melanoma skin cancer, dementia, and Parkinson’s disease, without delirium as there were only 2 samples with a history of delirium at baseline in the test set sample). P_adj_ (Full): p-values adjusted for multiple testing (tests based on age-adjusted, partially adjusted, and fully adjusted models for all-cause mortality and incident diseases). Disease/all-cause mortality highlighted with asterisk (*) if p_adj_ <0.05.

### 3.5 PAC versus other BA measures in associations with all-cause mortality and incident diseases

PAC outperformed PhenoAge, BioAge, and short LTL for most outcomes, showing the strongest associations with all-cause mortality, heart failure, pneumonia, delirium, COPD, dementia, lung cancer, myocardial infarction, osteoporosis, Parkinson’s disease, any cancer, and colorectal cancer (**Figure 2)**. In contrast, the associations with type 2 diabetes and chronic kidney disease were strongest with PhenoAge. BioAge showed the strongest associations with stroke and hypertension (**Figure 2**). Similar associations were observed in the age-adjusted and partially adjusted models (**Figures S5 and S6)**. Sensitivity analyses, including only disease-free participants at baseline, showed similar associations of BA acceleration with all-cause mortality and incident diseases, highlighting the robustness of our findings (**Figure 3**). Interestingly, the associations of PAC proteomic age acceleration with lung cancer and dementia were stronger in the disease-free participants than in the test set sample.

**Figure 2.**
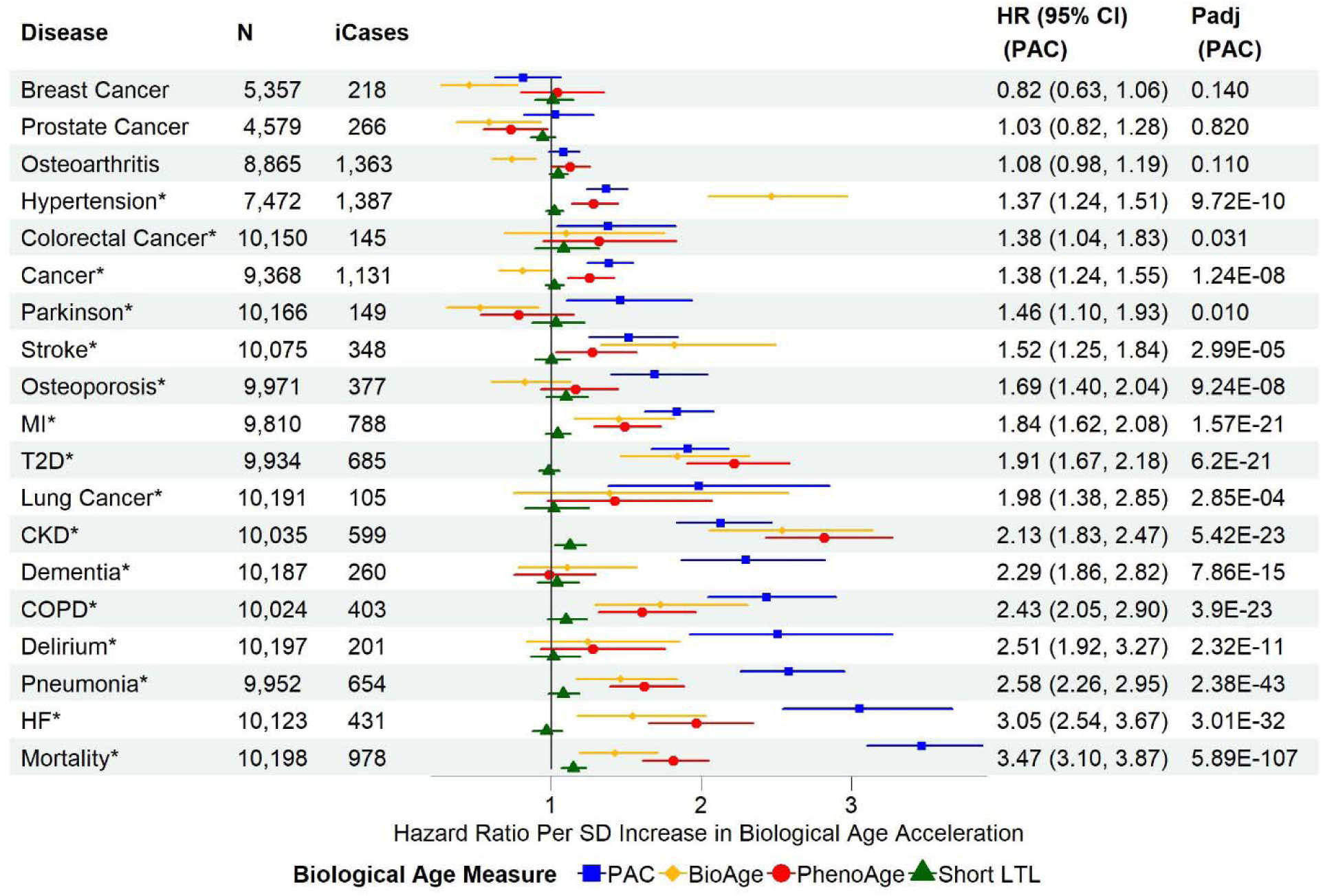
Associations of biological age acceleration based on different biological age measures with all-cause mortality and incident diseases in the test set sample using the fully adjusted models. **SD**: standard deviation of each measure after the inverse normal transformation in the combined training and test set to convert the original measurements to z-scores (approximately 1 across measures). **MI**: myocardial infarction; **T2D**: type 2 diabetes; **COPD**: chronic obstructive pulmonary disease; **CKD**: chronic kidney disease; **HF**: heart failure. **N**: sample size with complete data for the fully adjusted models of PAC, BioAge, PhenoAge, and short LTL, after excluding participants diagnosed with the disease at or prior to baseline. **ICases**: number of incident cases of N samples. Cox regression model for all-cause mortality and Fine-Gray subdistribution hazard models to account for the effect of death for the risk of incident diseases. The full covariate adjustment included chronological age, sex, ethnicity, education, Townsend deprivation index, smoking status, body mass index, and pre-existing diseases (hypertension, myocardial infarction, heart failure, stroke, type 2 diabetes, COPD, pneumonia, chronic kidney disease, any cancer excluding non-melanoma skin cancer, dementia, and Parkinson’s disease). Delirium was not included as there were only two samples with a history of delirium at baseline. **P_adj_**: p-values adjusted for multiple testing per BA measure (tests based on age-adjusted, partially adjusted, and fully adjusted models for all-cause mortality and incident diseases)

**Figure 3.**
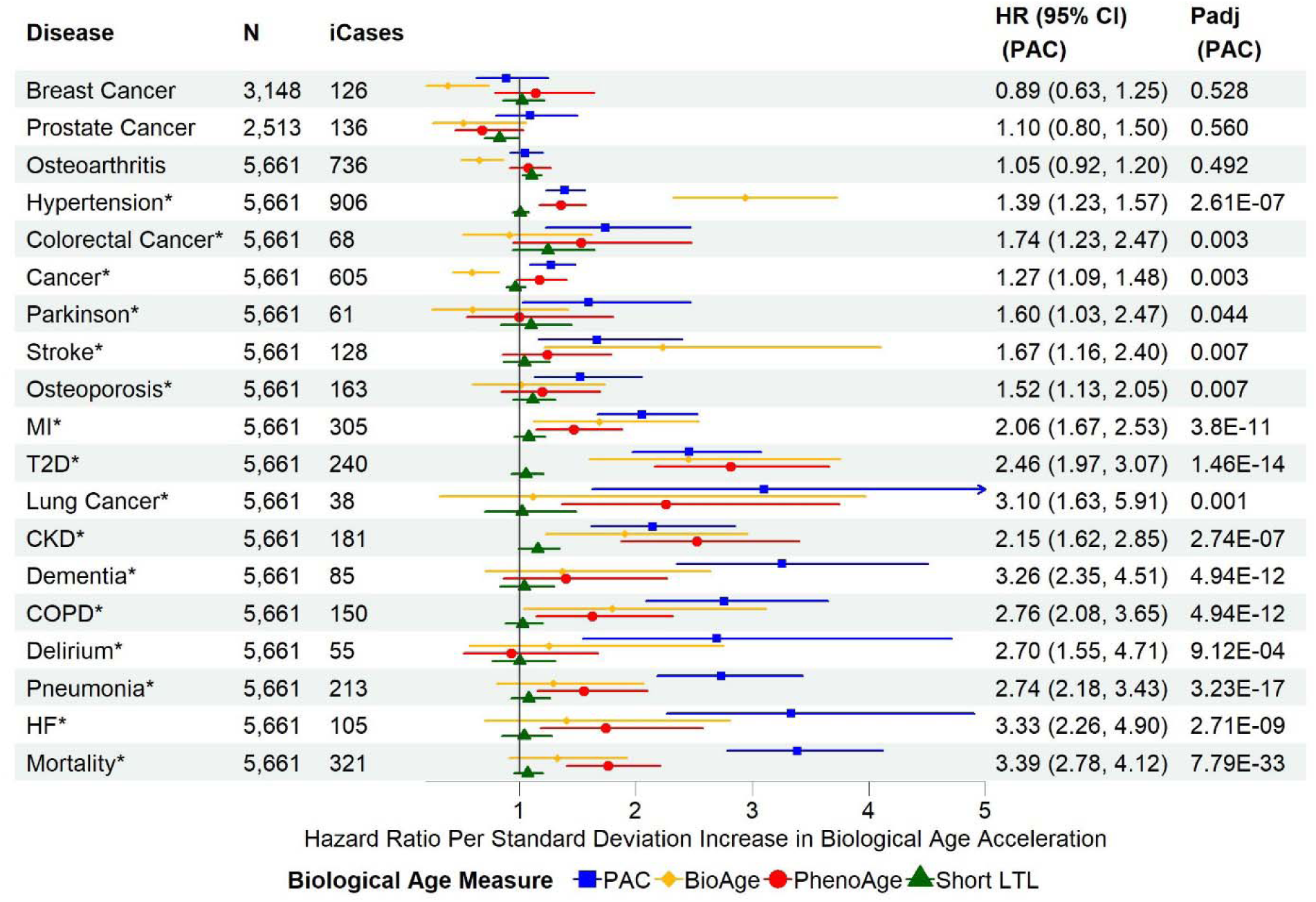
Associations of biological age acceleration based on different biological age measures with all-cause mortality and incident diseases using disease-free participants in the combined training and test set, and fully adjusted models. **SD**: standard deviation of each measure after the inverse normal transformation in the combined training and test set to convert the original measurements to z-scores (approximately 1 across measures). **MI**: myocardial infarction; **T2D**: type 2 diabetes; **COPD**: chronic obstructive pulmonary disease; **CKD**: chronic kidney disease; HF: heart failure. **N**: sample size with complete data for partially adjusted models of PAC, BioAge, PhenoAge, and short LTL, after excluding participants diagnosed with any of the diseases at or prior to baseline. **ICases**: number of incident cases of N samples. Cox regression model for all-cause mortality and Fine-Gray subdistribution hazard models to account for the effect of death on the risk for incident diseases. The partial covariate adjustment included chronological age, sex, ethnicity, education, Townsend deprivation index, smoking status, and body mass index. **Padj**: p-values adjusted for multiple testing per biological age measure (n=19)

### 3.6 PAC versus other BA measures in predictions for all-cause mortality and incident diseases

Using the test set data only, we compared the C-statistics for all-cause mortality and incident diseases of four models: 1) chronological age only (**M-Age**), 2) PAC proteomic age only (**M-PAC**), 3) BioAge only (**M-BioAge**), and 4) PhenoAge only (**M-PhenoAge**). M-PAC outperformed other models based on C-statistics, particularly all-cause mortality, COPD, pneumonia, and heart failure (**Figure 4**). The M-PAC C-statistics for dementia and delirium were the highest across diseases and models but not significantly different from those of M-Age (**Figure 4**). Models with multiple BA measures showed minimally improved C statistics (**Figure S7**).

**Figure 4.**
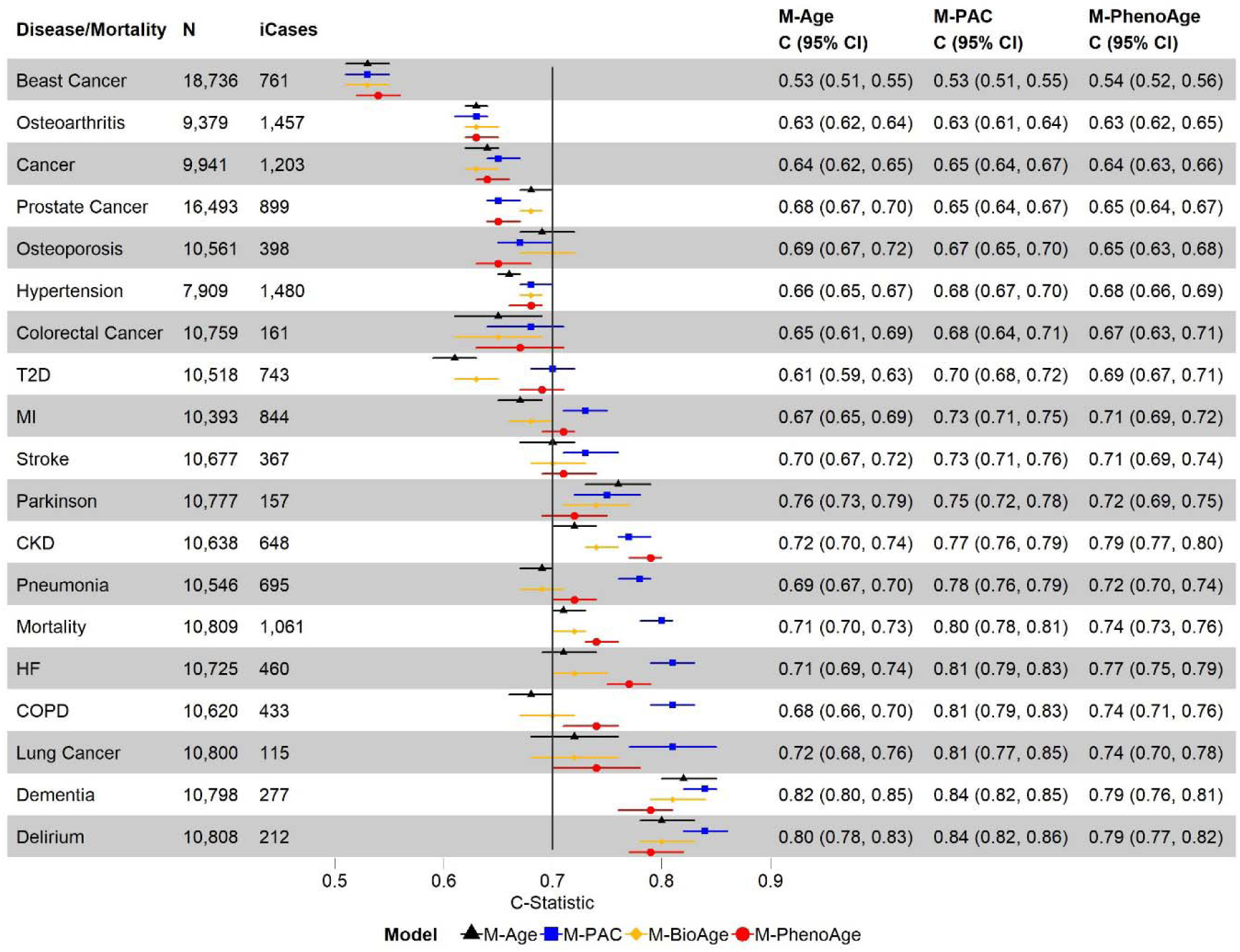
Discriminative power of biological age based on different measures for all-cause mortality and incident diseases using Cox models and the test set data: 1) model with chronological age only (M-Age), 2) model with PAC proteomic age only (M-PAC), 3) model with BioAge only (M-BioAge), and 4) model with PhenoAge only (M-PhenoAge). **MI**: myocardial infarction; **T2D**: type 2 diabetes; **COPD**: chronic obstructive pulmonary disease; **CKD**: chronic kidney disease; **HF**: heart failure. **N**: sample size with complete data for chronological age, PAC proteomic age, BioAge, and PhenoAge, after excluding participants diagnosed with the disease at or prior to baseline. **iCases**: number of incident cases of N samples.

### 3.7 Functional analysis

A total of 1,001 significant proteins coded by 1,008 genes were selected for significant associations with PAC proteomic age deviation to initiate a functional analysis by FUMA (**Table S7**). Genes associated with PAC proteomic age deviation were enriched in 25 hallmark gene sets (Bonferroni-corrected p<0.05) (**Figure 5**, **Table S11**). These hallmark gene sets include a wide range of biological processes and signaling pathways, particularly epithelial-mesenchymal transition, coagulation, inflammatory response, allograft rejection, IL-6-JAK-STAT3 signaling, complement, and IL2-STAT5 signaling (**Figure 5**). Additionally, genes associated with PAC proteomic age deviation were overrepresented in the differentially expressed genes in multiple tissues, topped by lung and adipose tissues (**Figure S6**).

**Figure 5.**
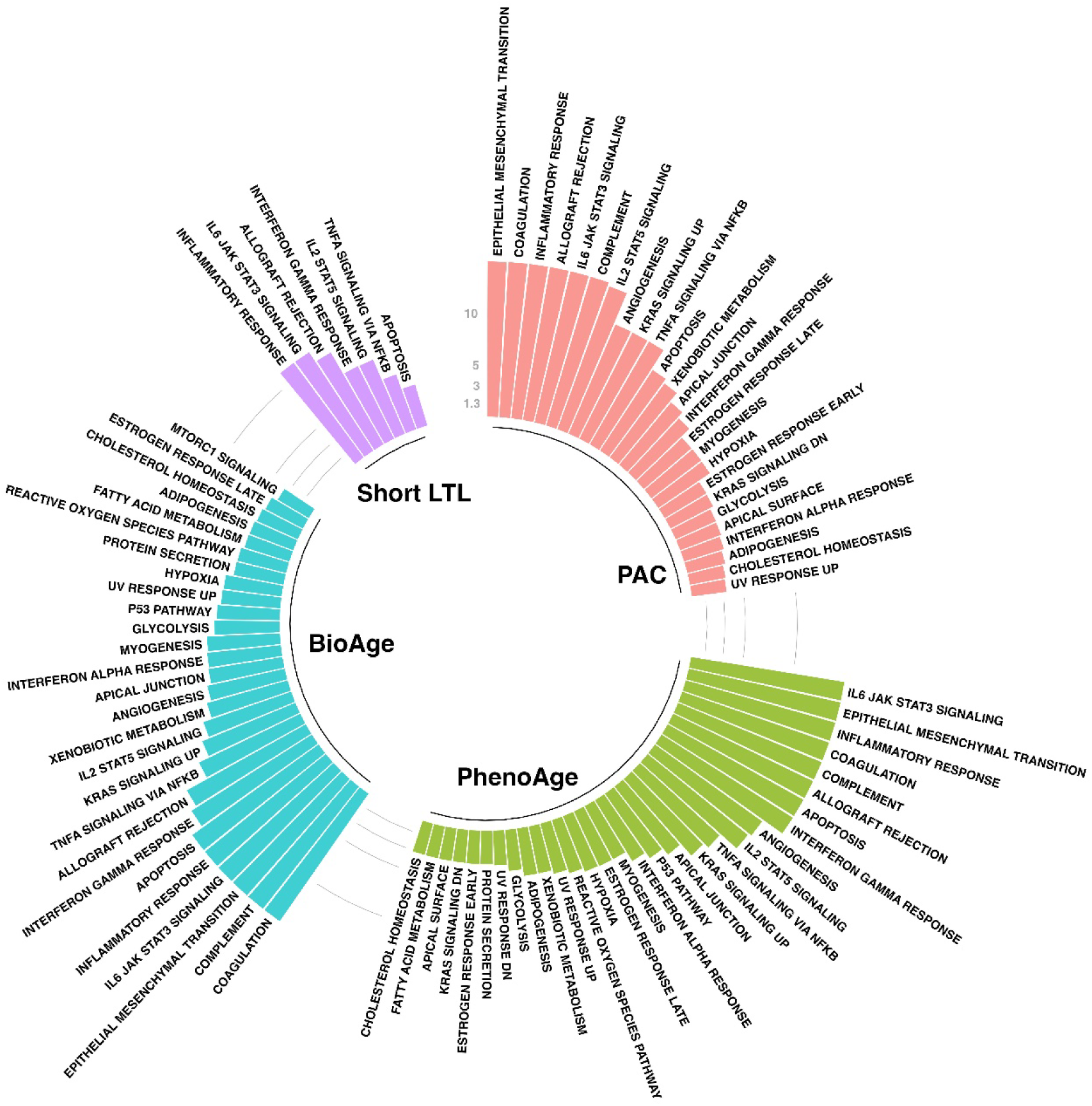
Hallmark gene sets enriched with genes associated with BA deviation based on different BA measures. The colored bars represent -log10(p) for different biological age measures after Bonferroni correction (n=50) and those greater than 15 are truncated at 15.

In contrast, proteins significantly associated with PhenoAge deviation (**Table S8**) were significantly enriched in 30 hallmark gene sets (**Table S12**) versus 27 hallmark gene sets with BioAge deviation (**Tables S9 and S13**), and seven hallmark gene sets with short LTL (**Tables S10 and S14**). Significant gene sets among Kyoto Encyclopedia of Genes and Genomes (KEGG) pathways or gene oncology (GO) resources, for example, were also reported in **Tables S11-S14**. These findings were consistent across BA measures (**Figures S8 and S9**), suggesting the presence of conserved biology underlying the aging processes.

## 4 Discussion

We developed a proteomic clock (PAC) for all-cause mortality as a surrogate of BA. PAC proteomic age acceleration was robustly associated with all-cause mortality and age-related diseases after controlling for sociodemographic, lifestyle factors, and pre-existing diseases, in the test set sample and disease-free participants. PAC proteomic age showed, in general, better performance in predicting all-cause mortality and incident diseases than chronological age and other BA measures. Proteins associated with PAC proteomic age deviation were enriched in various hallmarks of biological aging, including immunoinflammatory responses, cellular senescence, extracellular matrix remodeling, cellular response to stressors, and vascular biology. Interestingly, these processes are conserved over multiple BA measures, suggesting that such biological abnormalities are conserved regardless of how BA measures are developed or trained. Overall, our findings demonstrate the robustness of this proteomic aging clock in predicting different adverse health outcomes and reflect the current understanding of the perturbations in multiple biological pathways in the aging process.

Previous studies evaluated the proteomic correlates of chronological aging using different proteomic platforms and assays (e.g., SOMAscan assays^®^ or mass spectrometry)(Tanaka et al. 2018; Sathyan et al. 2020; Sayed et al. 2021). Proteins associated with chronological age showed significant associations with age-related clinical outcomes: walking speed, grip strength, frailty, multimorbidity, and all-cause mortality. A recent preprint (Argentieri et al. 2023) used a smaller protein panel (n=1,459) from the initial release of UKB-PPP (n=31,581) and data from China Kadoorie Biobank (n=1,418) to train a proteomic clock (ProtAge) to predict chronological age. The trained ProtAge was associated with all-cause mortality and several diseases, e.g., Alzheimer’s disease. However, there are marked differences between the two studies. First, we used a larger set of proteins to train a proteomic clock (2,920 vs. 1,459 proteins), providing a broader coverage of the human proteome. Second, we trained a proteomic clock to predict mortality instead of chronological age, a shift from a “first-generation” to a “second-generation” clock. To the best of our knowledge, PAC is the first proteomic aging clock developed for all-cause mortality risk as a surrogate of BA, using the largest dataset of proteins and individuals in the world. Lastly, we went beyond the ProtAge analyses and also reported C-statistics to show the predictive power of PAC versus other BA measures. Although the PAC and ProtAge are not directly comparable, PAC consistently showed high predictive power for multiple aging outcomes. Overall, our results expand previous findings by showing that PAC age acceleration strongly predicts all-cause mortality and several incident disease outcomes, with a follow-up exceeding a decade and a substantial sample size to ensure adequate statistical power.

PAC proteomic age acceleration showed the strongest associations with mortality risk and several disease outcomes (e.g., heart failure, pneumonia, delirium, COPD, and dementia). On the other hand, BioAge showed the strongest association with hypertension and stroke, whereas PhenoAge showed the strongest associations with type 2 diabetes and chronic kidney disease. This pattern of associations remained similar among individuals who had no medical comorbidity at baseline, except the associations of PAC proteomic age acceleration with lung cancer and dementia became stronger. These findings suggest that different BA measures may be implemented depending on the study context or the outcomes of interest and that PAC is particularly valuable in identifying high-risk individuals years before the earliest manifestations of chronic conditions.

We found that genes associated with proteomic age deviation are enriched in various hallmarks of biological aging, including immunoinflammatory responses, cellular senescence, extracellular matrix remodeling, cellular response to stressors, and vascular biology. Additionally, several hallmark gene sets are conserved across BA measures, including inflammatory response, allograft rejection, IL-6-JAK-STAT3 signaling, IL2-STAT5 signaling, TNG alpha signaling via NF-κB, and apoptosis. Our findings suggest that regardless of the BA measures used, our findings indicate consistent manifestations of biological processes and pathways in BA acceleration. The multifaced biological influence on aging phenotypes reinforces the potential for geroscience-guided interventions to target multiple age-related outcomes.

It is crucial to validate a biomarker of aging by comparing it with alternative measures using external cohorts. While we have validated the PAC, including a comparison with other BA measures using an internal independent sample, there has yet to be any external validation or incorporation of previous proteome-based measures. Other cohorts with proteomic data, like the Cardiovascular Health Study, Atherosclerosis Risk in Communities (ARIC) study, and Framingham Heart Study (FHS), use different assays (e.g., SOMASCAN assays) with significant variations in the protein coverage (Ngo et al. 2016; Norby et al. 2021; Austin et al. 2022). These technical differences across platforms (Eldjarn et al. 2023) prevent the interchangeable use of assays for external model validation and a direct comparison of results. For example, Tanaka et al. (Tanaka et al. 2018) and Sathyan et al. (Sathyan et al. 2020) developed predictors for chronological age using elastic net regression models. Their models require 76 and 162 proteins to estimate biological ages accurately. However, only a subset of these proteins is available in the UK Biobank (59 and 76, respectively), thus preventing the derivation of their measures in the UK Biobank. Additional efforts are warranted to reconcile these differences and assess the robustness and generalizability of such predictors across different datasets and populations (Moqri et al. 2024).

This study has additional limitations that need to be considered when interpreting our findings. *First*, we did not exclude deaths unrelated to biological aging, such as those resulting from accidents. However, such incidents are rare in the UKB cohort and unlikely to impact our findings significantly. *Second*, we could not compare PAC with commonly used epigenetic clocks since UKB does not have data on DNA methylation. However, PhenoAge was used to train DNAm PhenoAge, thus providing an indirect comparison between PAC and DNAm PhenoAge clock. *Third*, while PAC is robustly associated with mortality and major chronic diseases, disease-specific (You et al. 2023) or organ-specific clocks (Oh et al. 2023; Sehgal et al. 2023) address heterogeneity within individuals and may thus be more favorable in certain contexts.

In conclusion, we have developed a novel proteomic aging clock termed PAC, which demonstrated robust associations and predictions for mortality and the onset of various diseases. The diverse hallmark gene sets linked with PAC proteomic age deviation highlight the potential efficacy of geroscience-guided interventions. Further validation is essential to ascertain the use of PAC across different settings.

## Abbreviations

BA: biological age
FDR: Benjamini-Hochberg false discovery rate
FUMA: Functional Mapping and Annotation of Genome-Wide Association Studies
GO: gene oncology
KEGG: Kyoto Encyclopedia of Genes and Genomes
LASSO: least absolute shrinkage and selection operator
LTL: leukocyte telomere length
NHS: UK National Health Service
NPX: normalized protein expression
PAC: proteomic aging clock
SASP: senescence-associated secretory phenotype
UKB: UK Biobank
UKB-PPP: UK Biobank Pharma Proteomics Project

## Author contributions

CLK and BSD designed the study. CLK, LCP, and JLA processed the data while CLK conducted the statistical analyses. The initial manuscript was drafted by CLK and BSD, with contributions from ZC and PL. All the authors reviewed and approved the final version.

## Data availability statement

Data access is granted upon application to the UK Biobank. The R code (Chen & Kuo n.d.) for computing PAC proteomic age can be obtained from the GitHub repository at https://github.com/kuo-lab-uchc/PAC.

## Acknowledgments

Access to UK Biobank data was granted under application no. 92647 “Research to Inform the Field of Precision Gerontology” (PI: Richard H. Fortinsky). This research used data assets made available by National Safe Haven as part of the Data and Connectivity National Core Study, led by Health Data Research UK in partnership with the Office for National Statistics and funded by UK Research and Innovation (research which commenced between 1 October 2020–31 March 2021 grant ref MC_PC_20029; 1 April 2021–30 September 2022 grant ref MC_PC_20058). This research also used data provided by patients and collected by the NHS as part of their care and support. Copyright © (year), NHS England. Re-used with the permission of the NHS England [and/or UK Biobank]. All rights reserved.

