## Supplementary Materials for "Proteomic aging clock (PAC) predicts age-related outcomes in middle-aged and older adults"

### **1 Supplementary Methods**

**2 Supplementary Figures**

**Supplementary Figure 1.** Spearman correlations between chronological age, PAC proteomic age, PhenoAge, BioAge, short LTL, physiological or cognitive measures, and a 49-item frailty index.

**Supplementary Figure 2.** Spearman correlations between chronological age, PAC proteomic age, PhenoAge, BioAge, short LTL, and disease-associated biomarkers.

**Supplementary Figure 3.** Associations of PAC proteomic age acceleration with all-cause mortality and incident diseases in males in the test set.

**Supplementary Figure 4.** Associations of PAC proteomic age acceleration with all-cause mortality and incident diseases in females in the test set.

**Supplementary Figure 5.** Associations of biological age acceleration based on different biological age measures with all-cause mortality and incident diseases in the test set sample using the age-adjusted models.

**Supplementary Figure 6.** Associations of biological age acceleration based on different biological age measures with all-cause mortality and incident diseases in the test set sample using the partially adjusted models.

**Supplementary Figure 7.** Discriminative power of biological age based on different measures for all-cause mortality and incident diseases using Cox models and the test set data.

**Supplementary Figure 8.** Tissue-specific gene expression analysis of genes associated with biological age deviation (p-values significant at the Bonferroni-corrected level 0.05/30 for 30 tissue types in red bars and others in blue bars).

**Supplementary Figure 9.** Hallmark gene sets linked with biological age acceleration across biological age measures. Those significant at the Bonferroni-corrected level of 5% (n=50) are highlighted in yellow.

**3 Supplementary Tables**

**Supplementary Table 1.** Included proteins in Cardiometabolic, Inflammation, Neurology, and Oncology panels.

**Supplementary Table 2.** UK Biobank field IDs to extract data for this project.

**Supplementary Table 3.** A descriptive summary of participant characteristics at baseline for the training and test samples separately versus other UK Biobank participants.

**Supplementary Table 4.** Selected proteins by a penalized Cox regression model for all-cause mortality to construct a proteomic aging clock.

**Supplementary Table 5.** Gompertz model for all-cause mortality including the selected 128 proteins and chronological age at baseline and that including chronological age at baseline only using the training set data.

**Supplementary Table 6.** Associations of PAC proteomic age acceleration with all-cause mortality and incident diseases in the test set.

**Supplementary Table 7.** Proteomic associations with PAC proteomic age adjusting for chronological age, sex, ethnicity, education, Townsend deprivation index, smoking status, body mass index, and pre-existing diseases.

**Supplementary Table 8.** Proteomic associations with PhenoAge adjusting for chronological age, sex, ethnicity, education, Townsend deprivation index, smoking status, body mass index, and pre-existing diseases.

**Supplementary Table 9.** Proteomic associations with BioAge adjusting for chronological age, sex, ethnicity, education, Townsend deprivation index, smoking status, body mass index, and pre-existing diseases.

**Supplementary Table 10.** Proteomic associations with short leukocyte telomere length adjusting for chronological age, sex, ethnicity, education, Townsend deprivation index, smoking status, body mass index, and pre-existing diseases.

**Supplementary Table 11.** Enriched pathways of genes significantly associated with PAC proteomic age acceleration, with the full covariate adjustment.

**Supplementary Table 12.** Enriched pathways of genes significantly associated with PhenoAge acceleration, with the full covariate adjustment.

**Supplementary Table 13.** Enriched pathways of genes significantly associated with BioAge acceleration, with the full covariate adjustment.

**Supplementary Table 14.** Enriched pathways of genes significantly associated with short leukocyte telomere length, with the full covariate adjustment.

**Supplementary Methods**

**Proteomic aging clock (PAC) development**

The PAC development involves multiple prediction models for mortality. Chronological age and 2,940 proteins after excluding three with a high missing rate were used as predictors. Due to the dependence among the predictors, a smaller set of them can achieve nearly optimal prediction accuracy. To minimize the number of predictors without sacrificing prediction accuracy, we applied a Least Absolute Shrinkage and Selection Operator (LASSO) penalized Cox regression model. In this model, *λ* is a tuning parameter controlling the strength of penalty applied to coefficients. A higher *λ* imposes greater penalty on coefficients, leading to increased shrinkage of coefficients towards zero.

λ was optimized using 10-fold cross validation. It was chosen as the largest value among those that yielded a deviance within one standard deviation of the minimum deviance. Deviance (D) is a measure of goodness of fit: the smaller the deviance, the better the fit. Mathematically, it can be expressed as D=-2×(*L* - *L*_saturated_), where *L* represents the maximum log-likelihood of the fitted model and *L*_saturated_ represents the log-likelihood of the saturated model. In the saturated model, there is a separate coefficient for each predictor per individual in the dataset. This model perfectly fits the data but is rarely used in practice due to its complexity and potential for overfitting. As a heuristic rule (i.e., one-standard-error rule) (Friedman et al. 2010), λ was chosen as the largest value among those that yielded a deviance within one standard deviation of the minimum deviance.

After *λ* was chosen, the predictors (in the vector $\boldsymbol{x}$) with a non-zero regression coefficient in the penalized Cox regression model were carried forward to fit a Gomperz model (Lee & Wang 2003), with the probability density function (pdf),

$f\left( t \right| a, b, \boldsymbol{\beta})=b\left( \boldsymbol{x} \right)exp(at-b(\boldsymbol{x})/a\left( e^{at}-1 \right))$,

where $t$ is the follow-up time, $\boldsymbol{\beta}$is a vector containing the regression coefficients associated with $\boldsymbol{x}$, $a$ and $b\left( x \right)=b\times exp(\boldsymbol{\beta}^{\boldsymbol{'}}\boldsymbol{x})$ are the shape and rate parameters of the Gompertz distribution. The shape parameter (a) influences the rate of change in the hazard function over time, while the scale parameter (*b*(x)) determines the baseline level of the hazard function. Hence, the cumulative distribution function (cdf) of the Gompertz model is given by

$F\left( T\leq t | a,b, \boldsymbol{\beta} \right)=1-exp\left\{ \left( -\frac{b\left( \boldsymbol{x} \right)}{a} \right)\times\left( e^{at}-1 \right) \right\}$.

$F(T_{j}\leq10|a, b,\boldsymbol{\beta})$ denotes the probability that the *j*-th individual will die within the next 10 years. Let$F\left( T\leq10 | a_{0}, b_{0}, \beta_{age} \right)=F\left( T\leq10 \right|a, b, \boldsymbol{\beta})$, where $F(T\leq10 | a_{0}, b_{0}, \beta_{age})$ is the cdf of the Gompertz model including chronological age only. Age as a variable on the left can be expressed as a function of chronological age on the right and proteins by

$PAC\left( \boldsymbol{x} \right)=\frac{1}{\beta_{age}}ln\left\{ \frac{a_{0}}{b_{0}(1-e^{{10a}_{0}})}\ln\left( 1-F\left( T\leq10 | a, b,\boldsymbol{\beta} \right) \right) \right\}$,

where the parameters ($a_{0}$, $b_{0}$, $\beta_{age}$, $a$, $b$, and $\boldsymbol{\beta}$) are estimated using the training set data. The age estimator is referred to as proteomic aging clock (PAC), developed to calculate the proteomic age based on the input data.

**PhenoAge and BioAge**

PhenoAge (Levine et al. 2018) was trained for all-cause mortality using 42 clinical biomarkers in the National Health and Nutrition Survey (NHANES) III cohort. Nine biomarkers (albumin, creatinine, glucose, C-reactive protein, lymphocyte percent, mean cell volume, red cell distribution width, alkaline phosphatase, and white blood cell count, with UKB data field IDs provided in **Supplementary Table 3**) and chronological age were selected by a Cox penalized regression model to construct PhenoAge in a Gompertz model. BioAge(Levine 2013) was constructed using the Klemera and Doubal’s method(Klemera & Doubal 2006), with chronological age and preselected aging-related biomarkers that showed moderate correlations with chronological age (Pearson correlations > 0.1) in NHANES III.

PhenoAge(Levine et al. 2018) and BioAge (Levine 2013) were derived in the UKB using baseline biomarker data. For each biomarker, extreme values exceeding the 99th percentile were replaced with the 99th percentile, while values falling below the 1st percentile were replaced with the 1st percentile. The PhenoAge equation in Levine et al.(Levine et al. 2018) was used to calculate PhenoAge. BioAge was calculated using the BioAge (Kwon & Belsky 2021) R package that constructed BioAge using the parameters associated with chronological age and nine biomarkers estimated in NHANES III without separating men and women: forced expiratory volume in the first second, blood urea nitrogen, albumin, alkaline phosphatase, C-reactive protein, creatinine, total cholesterol, glycated hemoglobin [HbA1c], and systolic blood pressure, with UKB data field IDs provided in **Supplementary Table 3**).

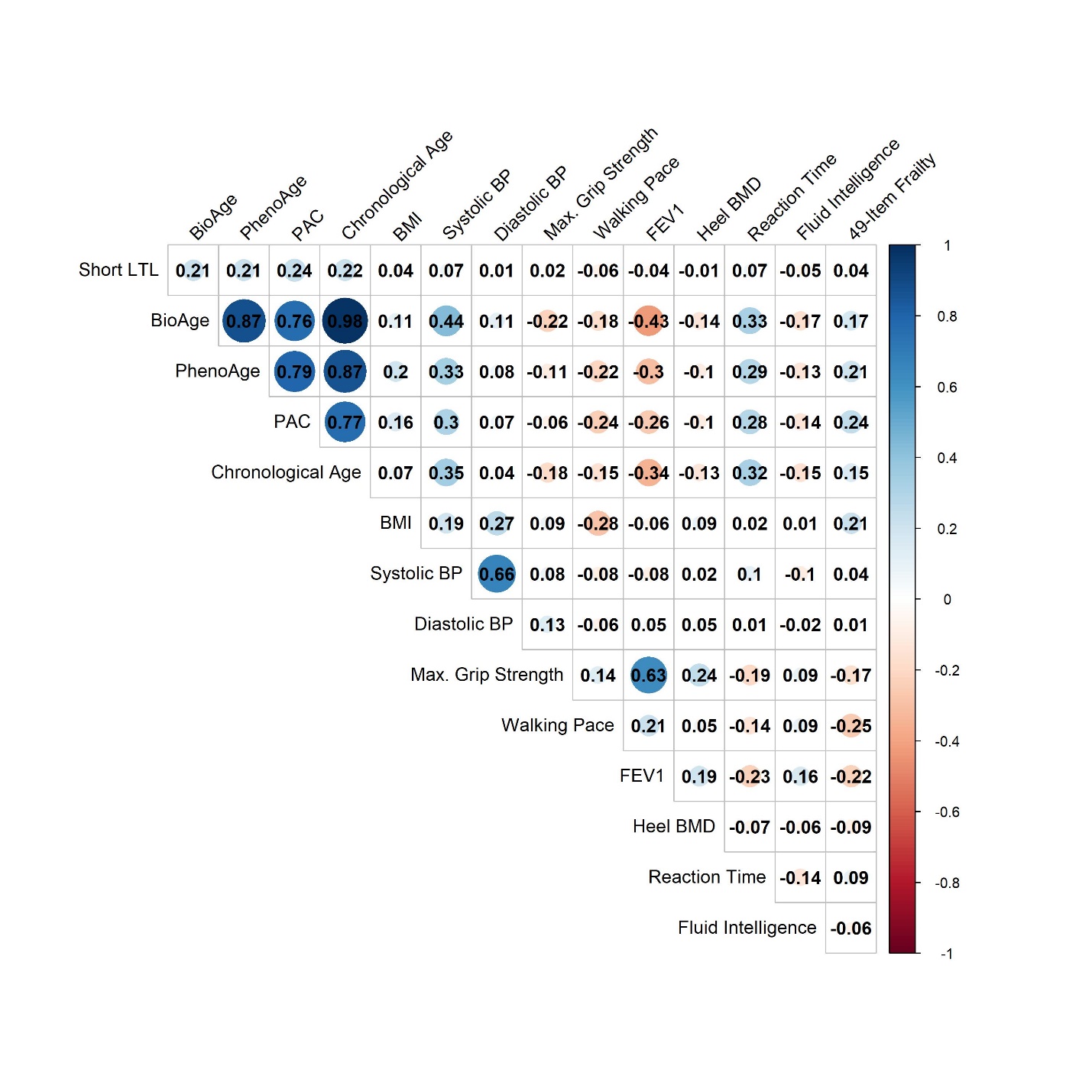

**Figure S1. Spearman correlations between chronological age, PAC proteomic age, PhenoAge, BioAge, short LTL, physiological or cognitive measures, and a 49-item frailty index.**

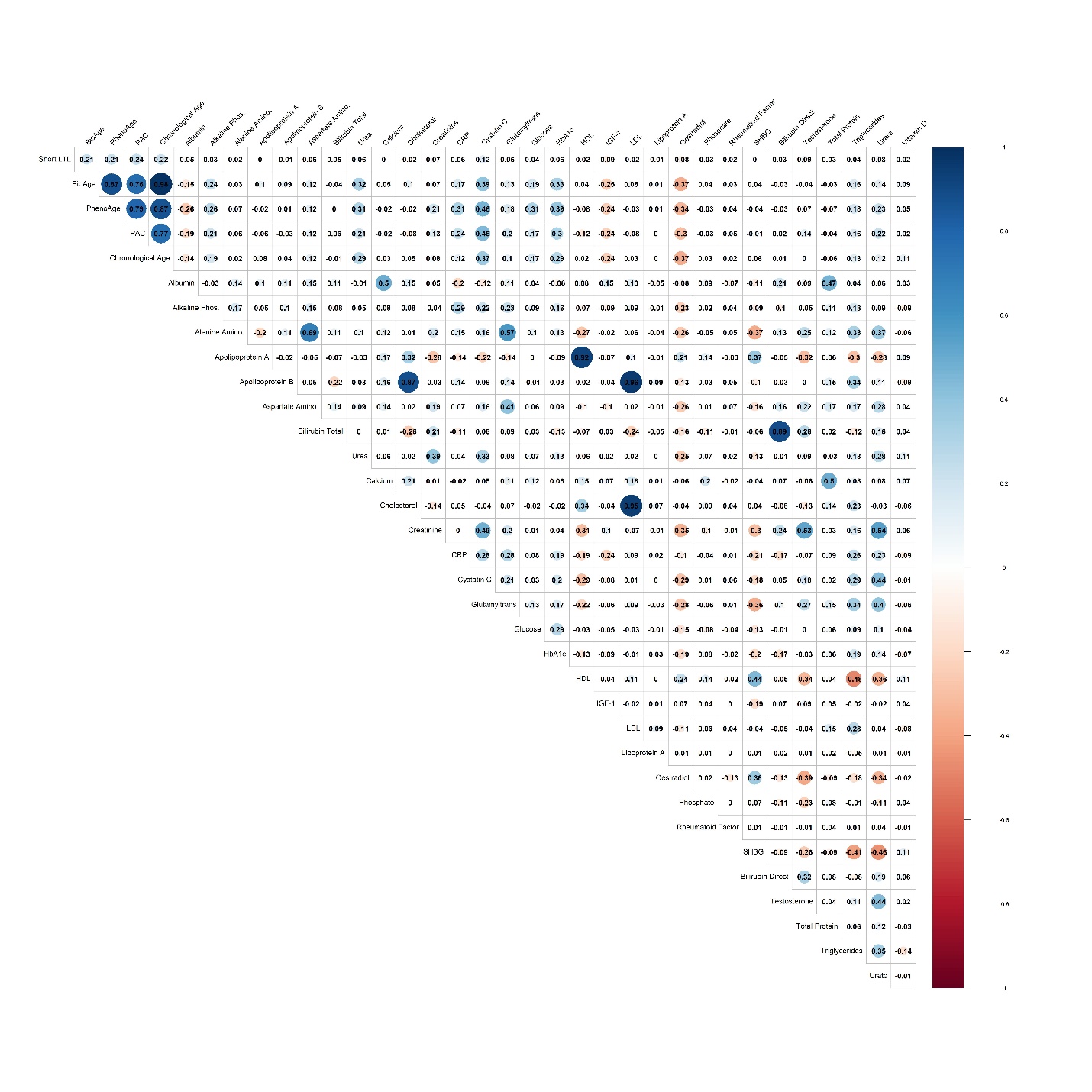

**Figure S2. Spearman correlations between chronological age, PAC proteomic age, PhenoAge, BioAge, short LTL, and disease-associated biomarkers.**

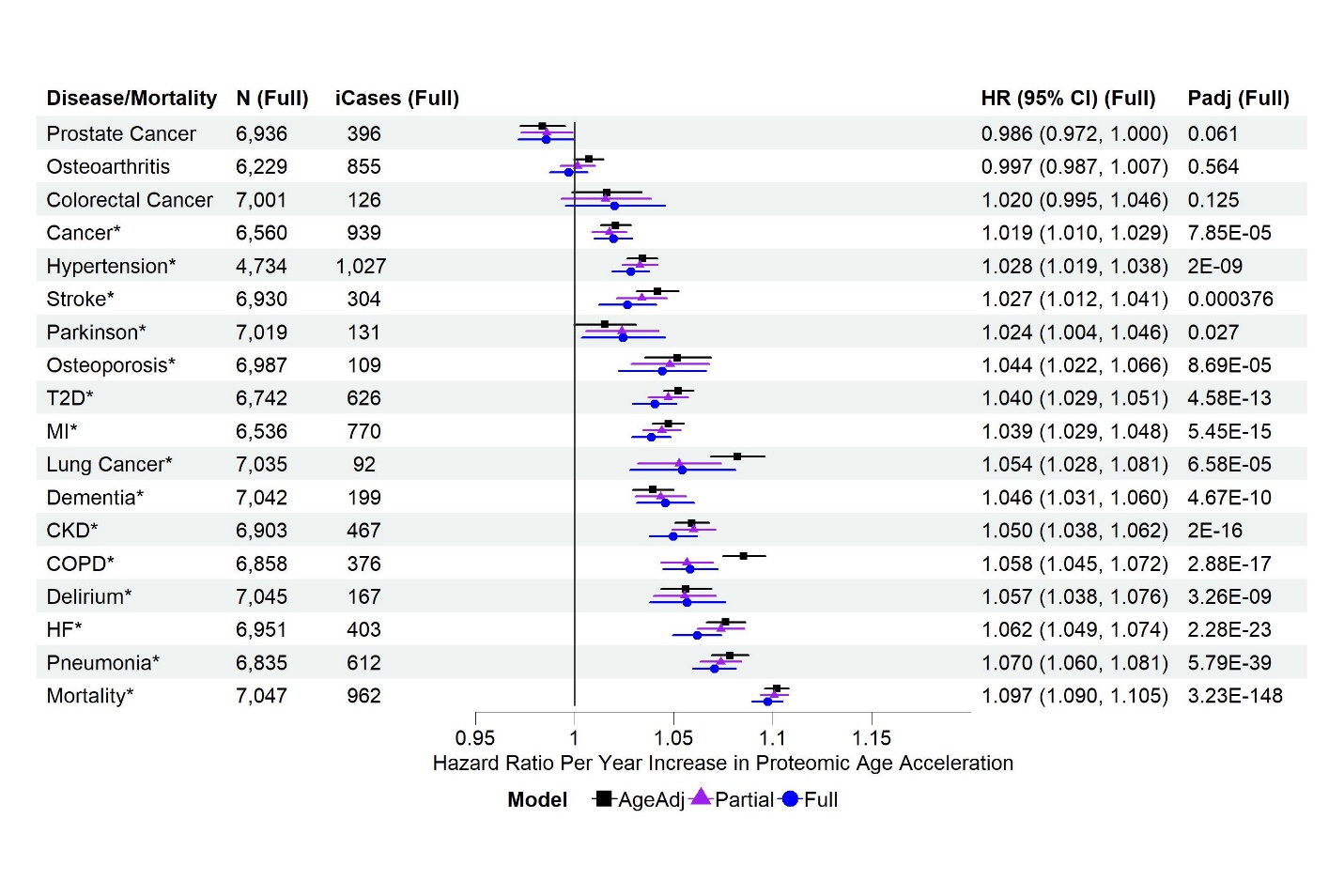

**Figure S3. Associations of PAC proteomic age acceleration with all-cause mortality and incident diseases in males in the test set. MI**: myocardial infarction; **T2D**: type 2 diabetes; **COPD**: chronic obstructive pulmonary disease; **CKD**: chronic kidney disease; **HF**: heart failure. **N (Full)**: sample size with complete data for the fully adjusted model, after excluding participants diagnosed with the disease at or prior to baseline. **iCases (Full)**: number of incident cases of N samples. Cox regression model for all-cause mortality and Fine-Gray sub-distribution hazard models to account for the effect of death on the risk for incident diseases, adjusting for different sets of covariates at baseline (age adjusted, partially adjusted, and fully adjusted models). **AgeAdj**: chronological age; **Partial**: chronological age, sex, ethnicity, education, Townsend deprivation index, smoking status, and body mass index; **Full**: covariates in the partially adjusted model, and pre-existing diseases (hypertension, myocardial infarction, heart failure, stroke, type 2 diabetes, COPD, pneumonia, chronic kidney disease, any cancer excluding non-melanoma skin cancer, dementia, and Parkinson’s disease, without delirium as there were only 2 samples with a history of delirium at baseline in the test set sample). **P_adj_ (Full)**: p-values adjusted for multiple testing (tests based on age adjusted, partially adjusted, and fully adjusted models for all-cause mortality and incident diseases). Disease/all-cause mortality highlighted with asterisk (*) if p_adj_ <0.05.

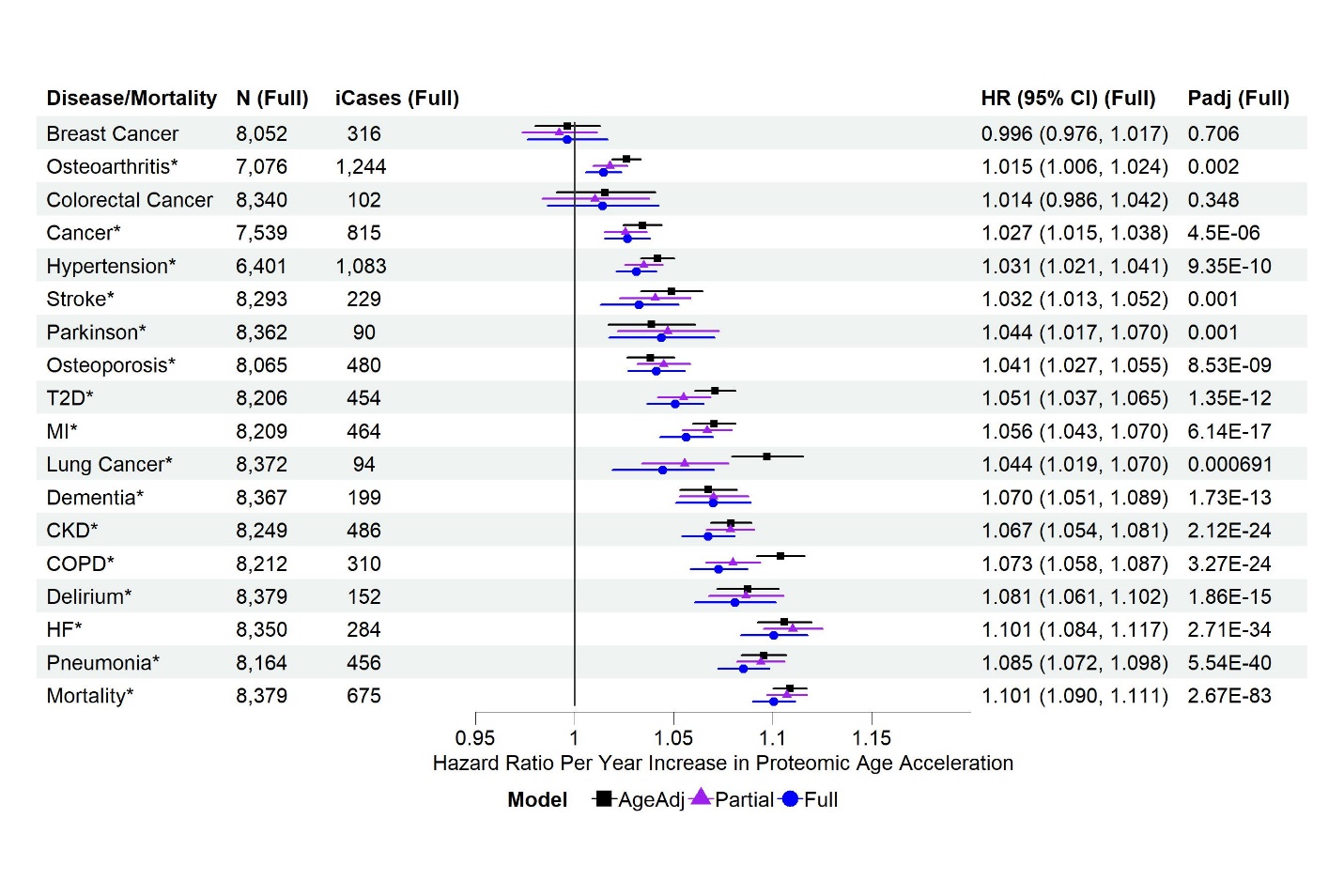

**Figure S4. Associations of PAC proteomic age acceleration with all-cause mortality and incident diseases in females in the test set. MI**: myocardial infarction; **T2D**: type 2 diabetes; **COPD**: chronic obstructive pulmonary disease; **CKD**: chronic kidney disease; **HF**: heart failure. **N (Full)**: sample size with complete data for the fully adjusted model, after excluding participants diagnosed with the disease at or prior to baseline. **iCases (Full)**: number of incident cases of N samples. Cox regression model for all-cause mortality and Fine-Gray sub-distribution hazard models to account for the effect of death on the risk for incident diseases, adjusting for different sets of covariates at baseline (age adjusted, partially adjusted, and fully adjusted models). **AgeAdj**: chronological age; **Partial**: chronological age, sex, ethnicity, education, Townsend deprivation index, smoking status, and body mass index; **Full**: covariates in the partially adjusted model, and pre-existing diseases (hypertension, myocardial infarction, heart failure, stroke, type 2 diabetes, COPD, pneumonia, chronic kidney disease, any cancer excluding non-melanoma skin cancer, dementia, and Parkinson’s disease, without delirium as there were only 2 samples with a history of delirium at baseline in the test set sample). **P_adj_ (Full)**: p-values adjusted for multiple testing (tests based on age adjusted, partially adjusted, and fully adjusted models for all-cause mortality and incident diseases). Disease/all-cause mortality highlighted with asterisk (*) if p_adj_ <0.05.

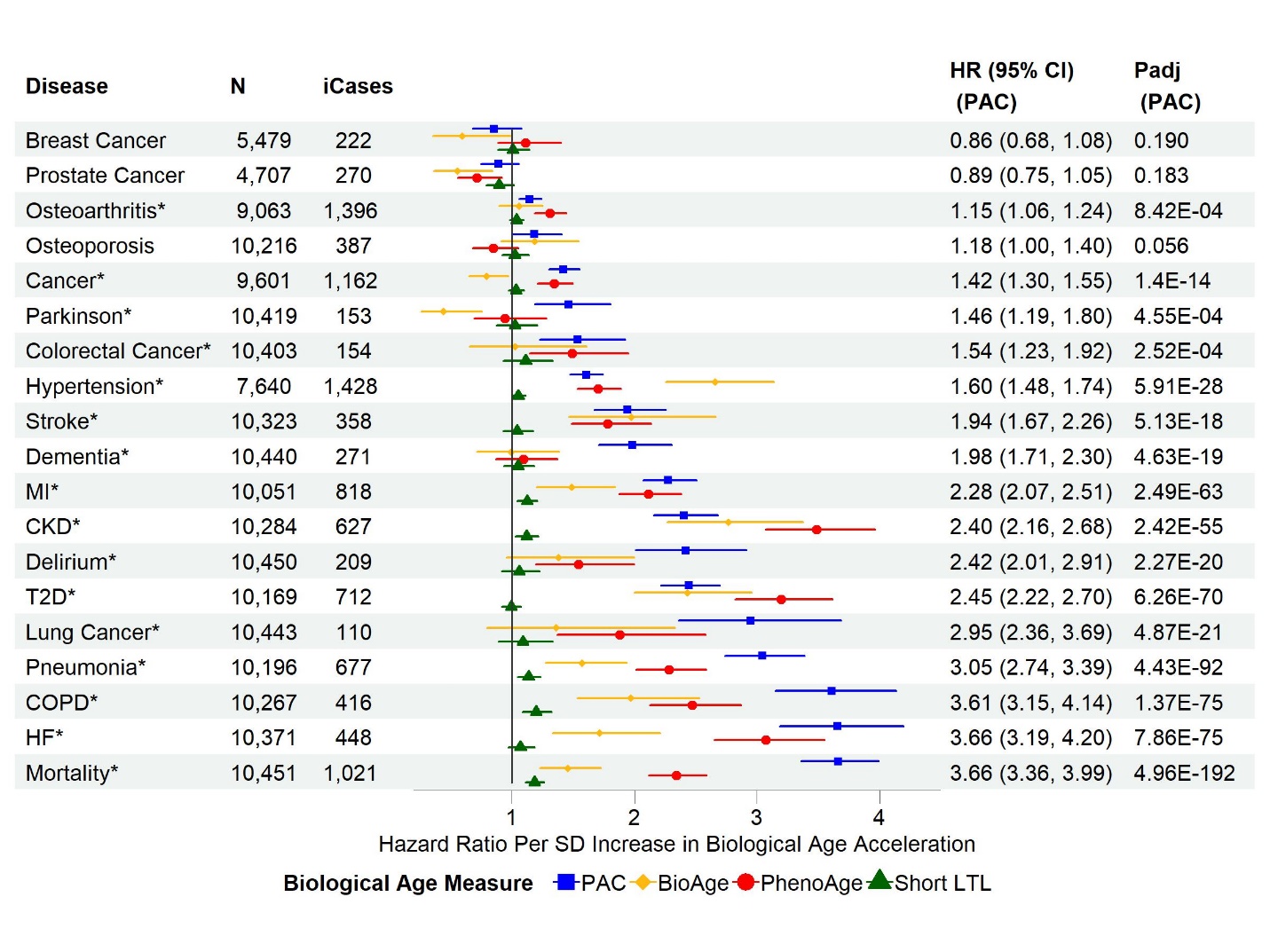

**Figure S5. Associations of biological age acceleration based on different biological age measures with all-cause mortality and incident diseases in the test set sample using the age-adjusted models.** **SD**: standard deviation of each measure after the inverse normal transformation in the combined training and test set to convert the original measurements to z-scores (approximately 1 across measures). **MI**: myocardial infarction; **T2D**: type 2 diabetes; **COPD**: chronic obstructive pulmonary disease; **CKD**: chronic kidney disease; **HF**: heart failure. **N**: sample size with complete data for the age-adjusted models of PAC, BioAge, PhenoAge, and short telomere length, after excluding participants diagnosed with the disease at or prior to baseline. **iCases**: number of incident cases of N samples. Cox regression model for all-cause mortality and Fine-Gray subdistribution hazard models to account for the all-cause mortality risk for incident diseases, with adjustment for chronological age. **P_adj_**: p-values adjusted for multiple testing per biological age measure (tests based on age-adjusted, partially adjusted, and fully adjusted models for all-cause mortality and incident diseases)

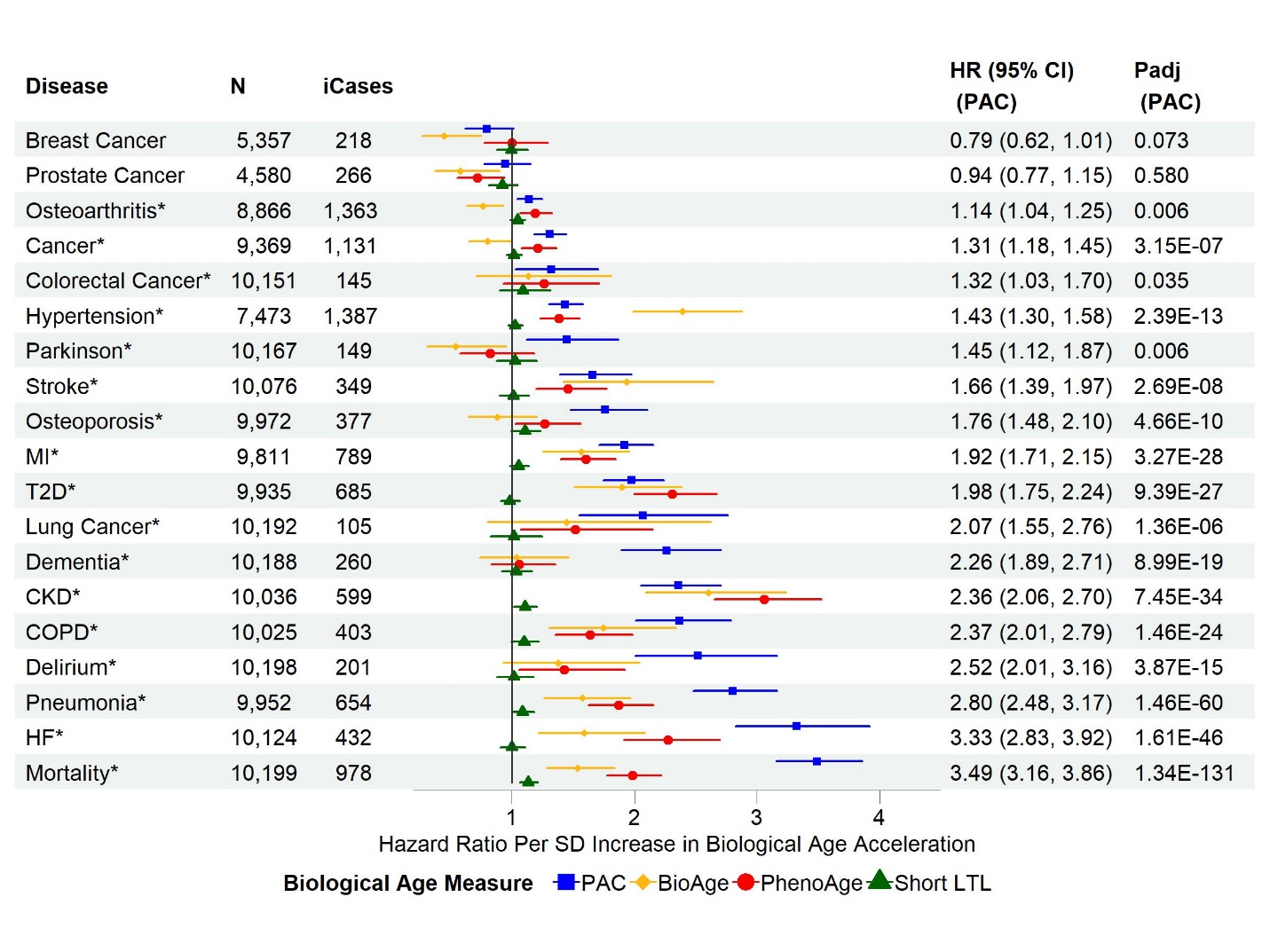

**Figure S6. Associations of biological age acceleration based on different biological age measures with all-cause mortality and incident diseases in the test set using the partially adjusted models.** **SD**: standard deviation of each measure after the inverse normal transformation in the combined training and test set to convert the original measurements to z-scores (approximately 1 across measures). **MI**: myocardial infarction; **T2D**: type 2 diabetes; **COPD**: chronic obstructive pulmonary disease; **CKD**: chronic kidney disease; **HF**: heart failure. **N**: sample size with complete data for the partially adjusted models of PAC, BioAge, PhenoAge, and short telomere length, after excluding participants diagnosed with the disease at or prior to baseline. **iCases**: number of incident cases of N samples. Cox regression model for all-cause mortality and Fine-Gray subdistribution hazard models to account for the all-cause mortality risk for incident diseases. The partial covariate adjustment included chronological age, sex, ethnicity, education, Townsend deprivation index, smoking status, and body mass index. **P_adj_**: p-values adjusted for multiple testing per biological age measure (tests based on age-adjusted, partially adjusted, and fully adjusted models for all-cause mortality and incident diseases)

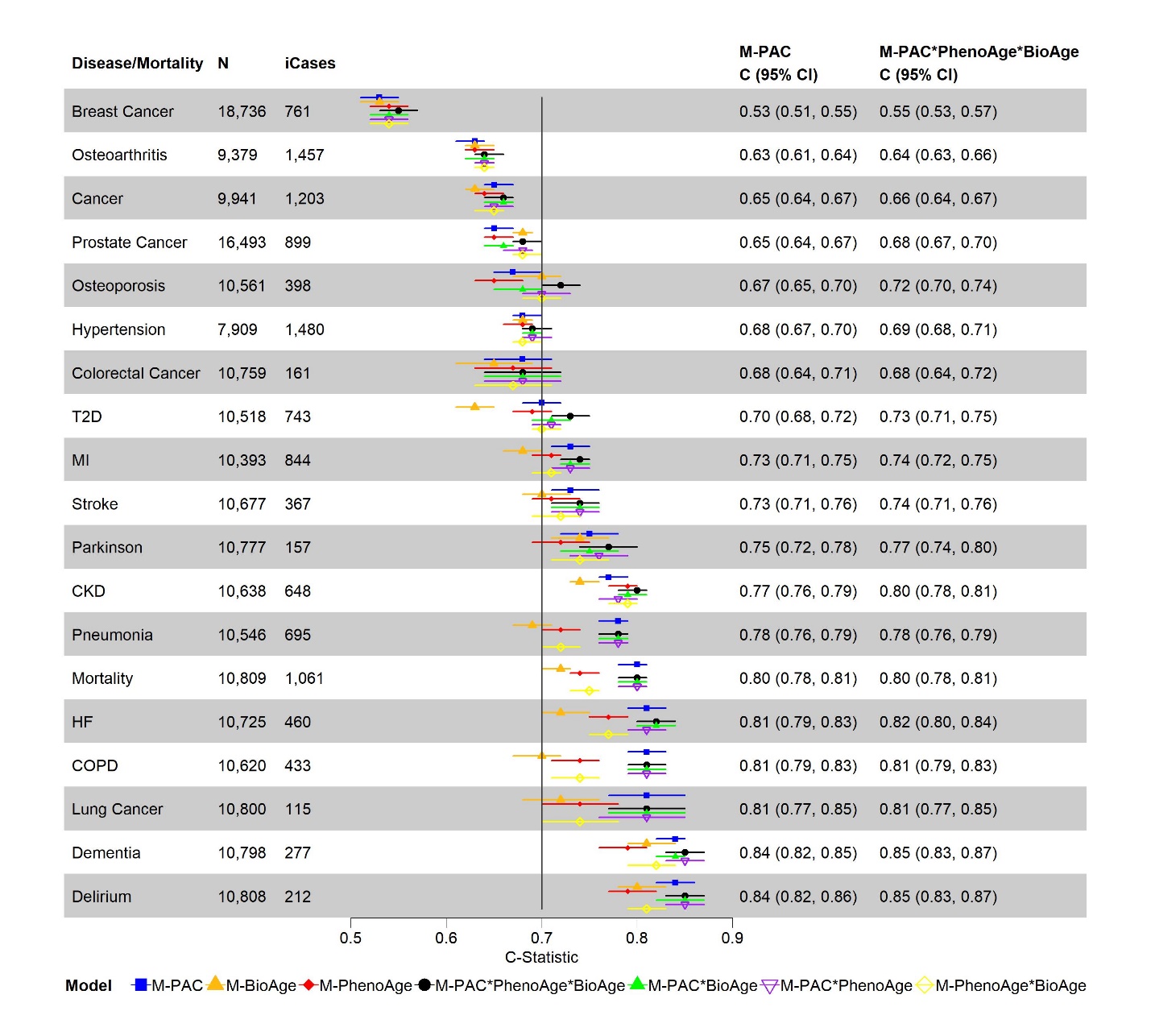

**Figure S7. Discriminative power of biological age based on different measures for all-cause mortality and incident diseases using Cox models and the test set data:** 1) model with PAC proteomic age only (M-PAC), 2) model with BioAge only (M-BioAge), 3) model with PhenoAge only (M-PhenoAge), 4) model with PAC proteomic age, BioAge, and PhenoAge, plus all the two-way and three-way interactions among PAC proteomic age, BioAge, and PhenoAge (M-PAC*PhenoAge*BioAge), 5) model with PAC proteomic age, BioAge, and the interaction between PAC proteomic age and BioAge (M-PAC*BioAge), 6) model with PAC proteomic age, PhenoAge, and the interaction between PAC proteomic age and PhenoAge (M-PAC*PhenoAge), 7) model with PhenoAge, BioAge, and the interaction between PhenoAge and BioAge (M-PhenoAge*BioAge). **MI**: myocardial infarction; **T2D**: type 2 diabetes; **COPD**: chronic obstructive pulmonary disease; **CKD**: chronic kidney disease; **HF**: heart failure. **N**: sample size with complete data of chronological age, PAC proteomic age, BioAge, and PhenoAge, after excluding participants diagnosed with the disease at or prior to baseline**. iCases**: number of incident cases of N samples.

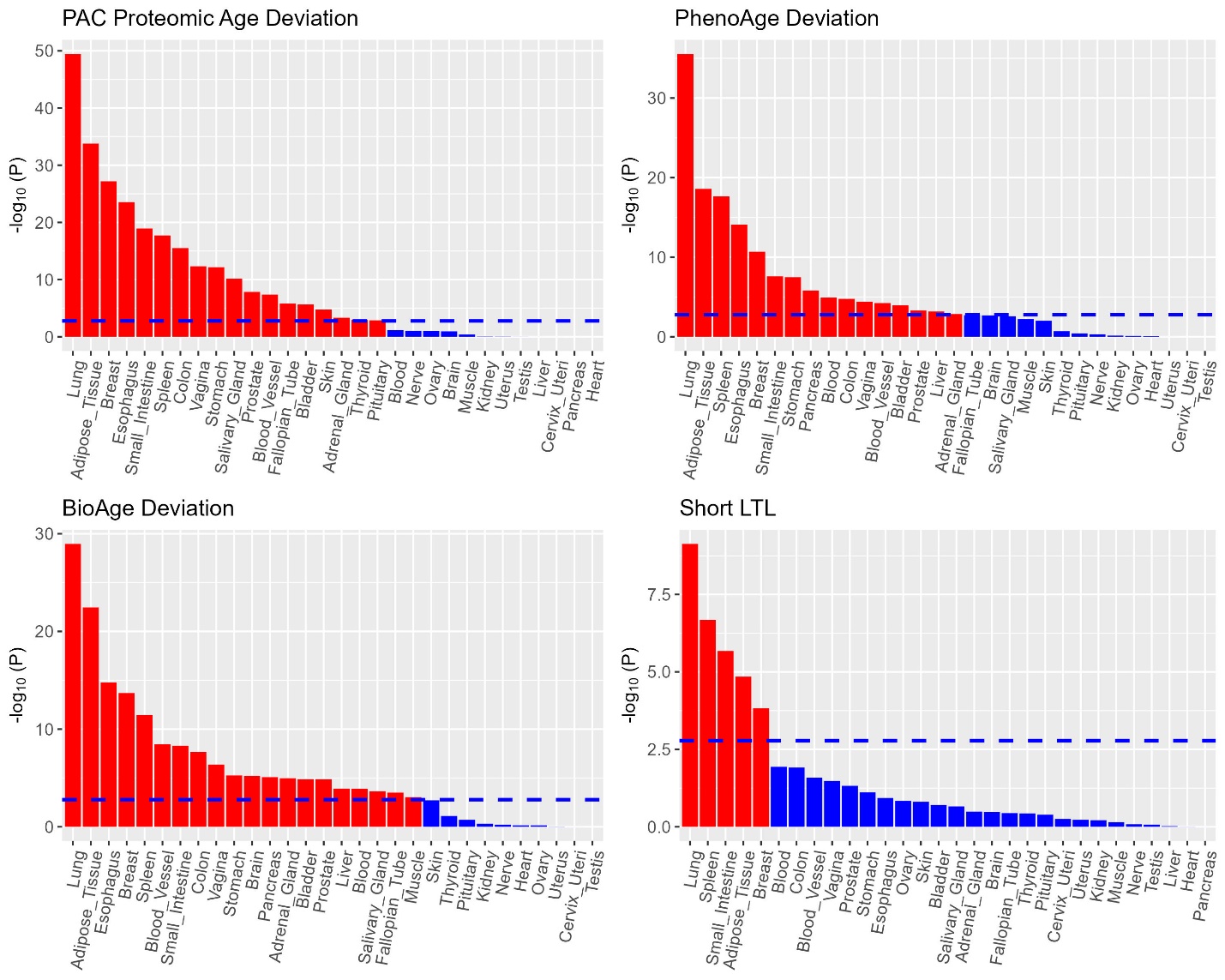

**Figure S8. Tissue-specific gene expression analysis of genes associated biological age deviation (p-values significant at the Bonferroni-corrected level 0.05/30 for 30 tissue types in red bars and others in blue bars)**

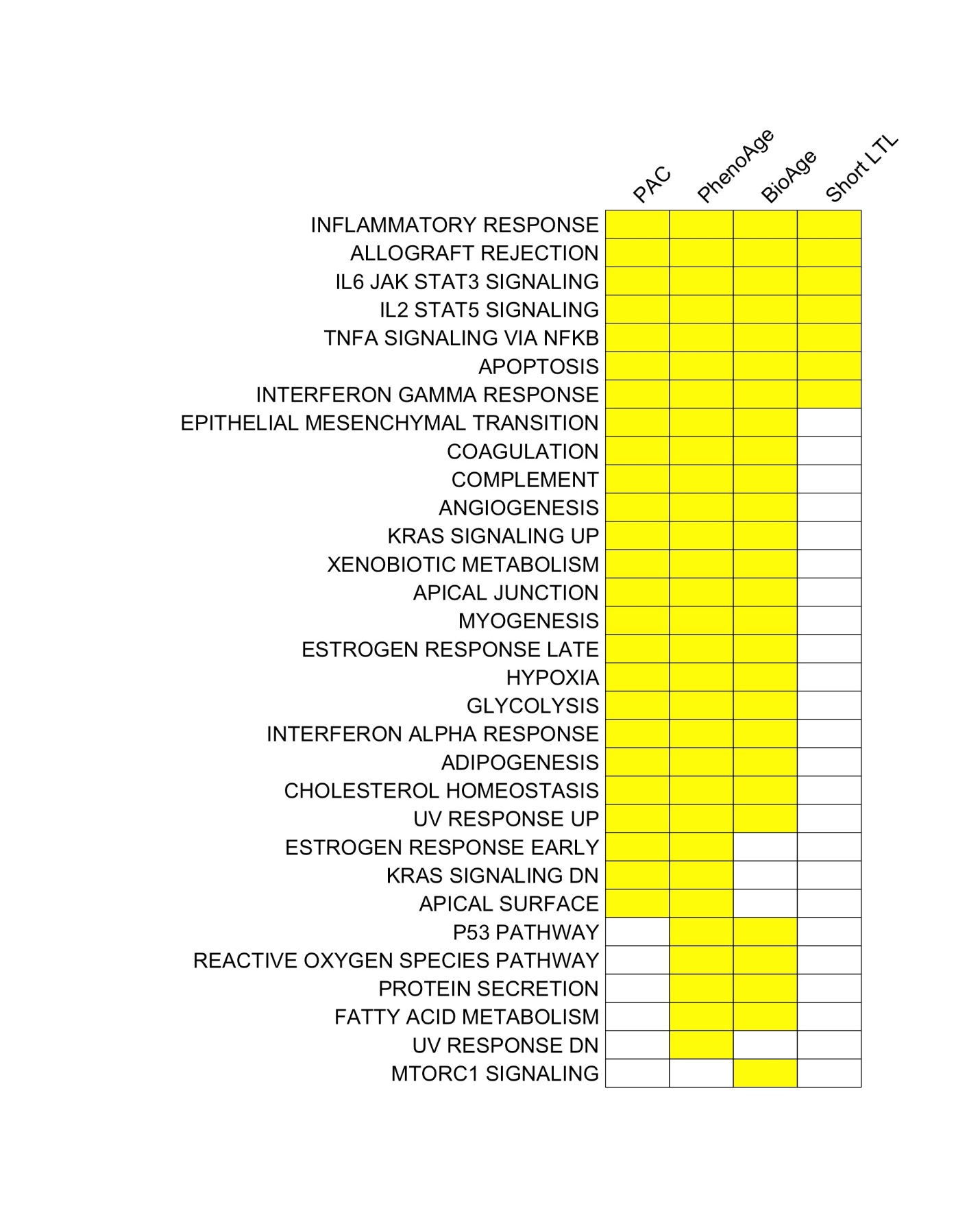

**Figure S9. Hallmark gene sets linked with biological age acceleration across biological age measures. Those significant at the Bonferroni-corrected level of 5% (n=50) are highlighted in yellow.**

**3 Supplementary Tables**

Download:

<https://docs.google.com/spreadsheets/d/1Qh-okXjyL3mBs9j645M0wFXakv1QpG82/edit?usp=drive_link&ouid=100617154994639126349&rtpof=true&sd=true>
